# Development of Rest–Activity Rhythms in Infancy and Their Disruption in Infantile Epileptic Spasms Syndrome

**DOI:** 10.64898/2026.08.07.26359346

**Authors:** Rima El Atrache, Saifina Karedia, Nandani Adhyapak, Anna Catherine Norman, Arindam Ghosh Mazumder, Danielle Schwartzenburg Takacs, Vaishnav Krishnan

## Abstract

**Background and Objectives:** In persons with epilepsy, seizure risk is tightly linked to the health of sleep and circadian rhythms. Rest-activity rhythms (RARs), derived from continuously worn activity monitors, can provide objective assessments of diurnal patterns of activity. Compared with healthy controls, adults with epilepsy have been shown to display weak and unstable RARs. In this study, we aimed to directly measure RARs in patients with infantile epileptic spasms syndrome (IESS), a potentially devastating developmental and epileptic encephalopathy. As a comparator, we similarly examined identically measured RARs from a cohort of healthy infants.

**Methods:** For this cross-sectional case-control comparison, we obtained multiday actograms in a sample of infants with IESS using ankle-worn Actiwatch-2 devices deployed during overnight follow-up EEG evaluations designed to assess initial treatment efficacy. Control actograms (similarly obtained via Actiwatch-2 devices) from the Rise & SHINE study (Sleep Health in Infancy and Early Childhood) were downloaded from the National Sleep Research Resource. We computed a series of parametric and non-parametric measures to depict the maturation of RARs over this developmental window and compared RARs from each IESS subject against up to 4 age-matched controls.

**Results:** In 891 actigraphy recordings obtained from 333 SHINE subjects, age-dependent increases in body length and weight were associated with progressive increases in RAR height (amplitude/mesor/M10), regularity (interdaily stability), entropy and fractal complexity, together with progressive declines in RAR fragmentation (intradaily variability). Compared with age-matched controls, multiday actograms from IESS subjects (n = 11, 9 males) displayed marked reductions in RAR height (amplitude/mesor/M10) and interdaily stability, together with reductions in entropy and fractal complexity.

**Conclusions:** During infancy, rest-activity rhythms display a stereotyped maturation in height, complexity and day to day consistency, revealing a developmental “growth curve” of RAR maturation. Severe RAR disruptions in infants with IESS may relate to the encephalopathy imposed by the underlying genetic/metabolic condition, structural lesion, and/or the psychomotor retardation imparted by antiseizure medications. Actigraphy recordings may offer a scalable, noninvasive approach to objectively and longitudinally assess circadian health in patients with IESS.

## Introduction

The first two years of life encompass a highly dynamic period of brain development, featuring marked increases in total brain volume and maturation. During this window, infants acquire progressively complex motor, sensory, and cognitive abilities^1^ driven by synaptogenesis, myelination and refinements in functional neural networks^2,3^. These changes occur in parallel with the establishment and maturation of circadian rhythms of rest, activity and sleep. In the late second trimester of human fetal development, randomly distributed bouts of spontaneous movement organize into ultradian rhythms of behavioral state cycling^4,5^. By the third trimester, a superimposed 24h rhythm of fetal heart rate, breathing movements and activity levels is entrained by maternal signals, including diurnal fluctuations in cortisol and body temperature^4,6^. Following birth, a transient return to ultradian rhythmicity is gradually overtaken by an increasingly organized circadian rest-activity pattern as the suprachiasmatic nucleus matures and integrates cues such as light, feeding and ambient temperature^3,7^. Sleep also transitions from 3-4h bouts distributed throughout the day, to longer bouts that are consolidated into a main nocturnal sleep period^8,9^. Disruptions to sleep and circadian health are often the earliest reported abnormalities in infants with neurodevelopmental disorders^10^. Separate from syndromic presentations, circadian and sleep abnormalities in otherwise neurotypically developing infants may serve as a risk factor for other childhood conditions, such as obesity^11,12^.

Quantitative assessments of 24h rest and activity rhythms (RARs) can be easily obtained using continuously worn activity monitors. From multiday recordings tallying activity “counts” (a compressed measure of acceleration over a 30 or 60s epoch^13^), such “actograms” can be used to derive a set of subject-specific metrics that quantify the strength, phase/timing and stability of RARs. Parametric approaches based on nonlinear regression (e.g., cosinor analysis) provide estimates of RAR height (amplitude, mesor) and timing (acrophase), together with measures of model fit (e.g., root mean squared error/RMSE)^14^. Nonparametric measures (e.g., intradaily variability (IV), interdaily stability (IS), etc.) directly quantify regularity, fragmentation, scaling behavior and complexity from time series structure without assuming a predefined waveform. Several large actigraphy phenotyping efforts employing hundreds to thousands of community-derived subjects (e.g., NHANES) have revealed how RAR metrics vary by age, sex and race/socioeconomic status^15-17^, as well as disease states, such as obesity^18^ and depression^19^. From prospective cohort studies of primarily adult subjects, specific RAR aberrations have also been linked to risk of future illness, including cardiovascular disease^20^ and cognitive decline^21^. The development and evolution of RARs in infants has received considerably less emphasis. A few longitudinal actigraphic assessments have described RAR maturation patterns qualitatively, employing smaller sample sizes (7-11 subjects)^9,22,23^. In the largest published study to date, Rojo-Wissar et al employed ankle actimeters to measure RAR metrics across 414 healthy infants at ages 3, 6 9 and 12 months of age. Compared with 3-month-old subjects, actograms from 6-12-month-olds displayed a progressive increase in amplitude, mesor and IS, together with decreases in IV^24^, indicating a gradual strengthening of circadian rhythmicity together with a consolidation rest and active periods.

In patients with epilepsy, where seizure risk is frequently accompanied by enduring changes in sleep and arousal patterns, cross-sectional case-control comparisons in adults have revealed fragmented and low amplitude RARs, featuring significantly increased IV, reduced IS^25,26^ and “M10” levels^25,26^ (mean activity counts accumulated during the 10 most active hours of the day). We previously reported similar findings in a cohort of adults with focal epilepsy^27^, and further showed that in those with epilepsy and intellectual disability, RARs are more severely weakened^28^, without a clear correlation to motor impairment. While the mechanisms underlying RAR disturbances remain incompletely understood, actigraphy-derived RAR measurements provide a noninvasive and holistic biomarker of diurnal behavior, agnostic to language function and amenable to repeated longitudinal testing.

In this study, we employ actigraphy to determine RAR aberrations in Infantile Epileptic Spasms Syndrome (IESS), a developmental and epileptic encephalopathy that typically emerges between 3-7 months of age^29^. The defining feature of IESS is infantile spasms: brief motor seizures that involve flexion, extension, or both of proximal and truncal muscles. Spasms tend to occur in clusters (often upon awakening), that can vary in intensity and duration^30^. IESS is etiologically diverse, spanning both acquired or congenital structural lesions and a wide range of genetic and metabolic etiologies^29,31^. Infantile spasms are treated with a combination of hormonal therapies (ACTH and/or prednisolone) and antiseizure medications (including vigabatrin). Although spasms generally resolve by five years of age, other seizure types may emerge and persist in many patients. IESS is often associated with developmental delay, autism, intellectual disability, cerebral palsy and pervasive epilepsy^32^. Early recognition and treatment improve outcomes^31^.

The links between IESS and circadian dysfunction are multifaceted. Spasms most commonly occur during sleep-wake transitions, typically between 9 am and noon^33^, and are associated with lower total daily sleep and diminished rapid eye movement (REM) sleep^34^. In a rat model of acquired epileptic spasms, reduced expression of circadian proteins (e.g., CLOCK, PER2) and the glucocorticoid receptor (GR-α) was alleviated by ACTH treatment^35,36^. ACTH itself lowers circulating levels of melatonin, which when exogenously supplemented, improved sleep quality in IESS patients without improving spasm frequency^37^. In this study, we aimed to characterize RAR abnormalities in infants with IESS using multi-day actigraphy. To contextualize our RAR measurements in IESS subjects, we take advantage of a publicly accessible annotated collection of actigraphy recordings from Rise and SHINE study (<u>S</u>leep <u>H</u>ealth in Infancy and <u>E</u>arly Childhood), where recordings were obtained from healthy infants between one month and 2 years of age^38-41^. Importantly, both SHINE and IESS cohorts wore the same FDA-cleared activity monitor (Actiwatch-2).

## Methods

### IESS Subjects

All study protocols were approved by the Baylor College of Medicine Institutional Review Board. We prospectively enrolled patients with newly diagnosed (EEG-confirmed) IESS at the Texas Children’s Hospital (Houston, TX) between May 2024 and May 2025. Following the initial diagnosis of infantile spasms, all patients were admitted for a scheduled follow-up vEEG evaluation approximately two weeks later, designed to assess treatment response. We obtained written informed consent from patient guardians prior to enrollment. Actiwatch-2 devices (Phillips Respironics) were deployed at the left or right ankle and configured to begin activity monitoring at 1700 on admission day, collecting activity counts in 60s epochs for the next 28 days. Families were instructed to obtain at least 10 full days of recording and then return the device in a prepaid envelope. From electronic health records, we extracted infant age, length and weight (at the time of admission), as well as results of brain imaging) and genetic diagnoses (when available). We reviewed associated raw EEGs and reports. Spasm onset times were manually annotated to minute-level activity recordings. IESS subjects and families were not provided any reimbursement for their participation.

### Control Subjects

Actigraphy recordings and demographic variables from the Rise and SHINE study were downloaded with permission from the National Sleep Research Resource (NSRR)^42^. SHINE enrolled mother-singleton dyads after delivery and prior to discharge from the newborn unit of the Massachusetts General Hospital (Boston, MA), between May 2016 and June 2018^41^. To be included, mothers had to be (i) fluent in either English or Spanish, (ii) at least 18 years of age, (iii) both the biological and birthing mother of the infant and (iv) without any significant health conditions. Included infants were healthy and full term, without known genetic disorders or congenital malformations that could impact sleep or growth^38^. Actiwatch-2 devices were deployed on the left ankle for a planned recording duration of 7 days^39-41^ during home visits that were conducted at 1, 6, 12 and 24 months of age (visits 1-4, respectively)^39^. Participants received a gift card at every visit^39^. SHINE demographic variables that we incorporated into our analyses included the age, sex, race and infant length/weight for each recording chosen. From all actograms available at NSRR, we excluded actograms with >10% missingness and ≤ 4 full days captured.

### Data analysis

Actograms from both cohorts were analyzed identically. Activity “counts” were tallied in 60s epochs (IESS) or 30s epochs (SHINE) that were subsequently down sampled to 60s epochs by addition. To calculate RAR metrics, we assembled contiguous whole days of recording (defined as midnight to midnight), after trimming “tail” segments. To provide a simple visualization that portrays gross differences in RAR height and morphology (Fig. A1/A2), we averaged minute-by-minute activity counts across all recording days and across participants to generate a group-level mean 24-hour actogram that was smoothed using a 100-neighbor moving average and a second order polynomial trend^43^. All subsequent analyses were performed on unsmoothed data. Heatmaps averaging the first six days of actigraphy are also shown^28^. To compute the relative power of ultradian and circadian periodicity (Fig. A1/A2), unsmoothed actograms were utilized to generate Lomb-Scargle periodograms (using MATLAB’s `plomb` function) over frequencies corresponding to period lengths between 0.5-40h (in 0.025h increments). Spectral power at each period length was normalized by dividing by the sum of total power across all tested periods^27,44^. For case-control comparisons (Figure 2), exact sex- and age-matching was not possible due to the demographic constraints of the SHINE dataset. For 9/11 IESS subjects, we selected 4 unique SHINE recordings from infants that were similar in age (+/-25 days). Two ∼10-month-old IESS subjects were together matched with a total of five age-matched SHINE recordings. In 4/11 IESS subjects that displayed persistent spasms during the follow up inpatient EEG evaluation, the timing of spasms was tallied using video EEG data (Fig. 3).

**Figure 1.**
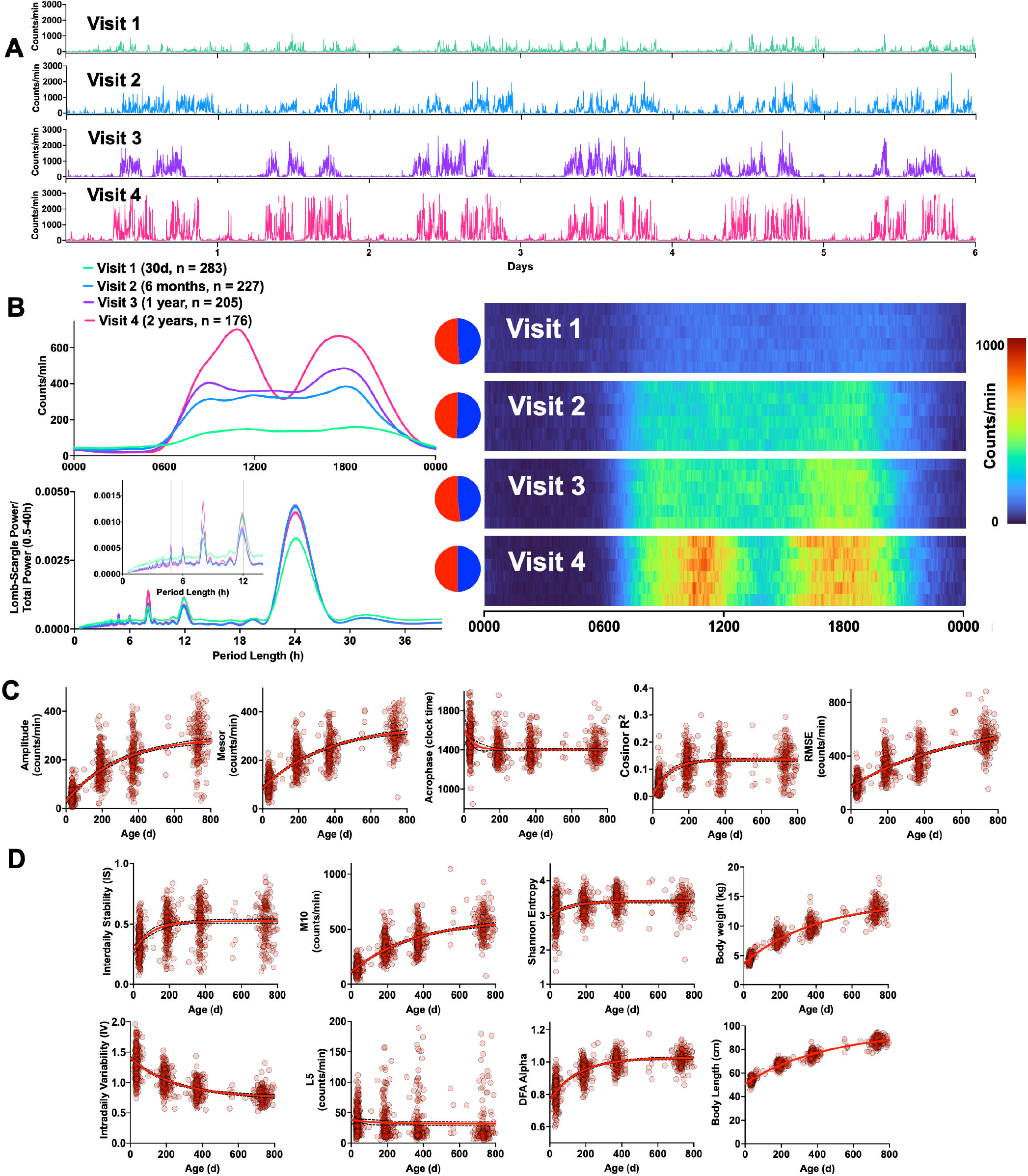
On the Maturation of Rest Activity Rhythms (RARs) During Infancy. A: Representative six-day raw actograms obtained from SHINE subjects during visits 1 (∼30d old), 2 (∼6 months old), 3 (∼1 year old) and 4 (∼2 years old). B: Smoothed and averaged actograms by visit (top left). Lomb-Scargle periodograms depicting the relative power of circadian and ultradian peaks averaged by SHINE visit (bottom left). RIGHT: Heatmaps depicting averaged (unsmoothed) actigraphy by SHINE visit for the first six full days of recording (stacked vertically). Pie charts reflect sex breakdown for each visit group (red: female, blue: male). C: Age-dependent increases in parametric endpoints (cosinor analysis), including amplitude, mesor, acrophase, R^2^ and root mean squared error (RMSE). D: Age-dependent changes in non-parametric endpoints, including IS, IV, M10, L5, Shannon entropy, alpha coefficient for detrended fluctuation analysis (DFA), body weight and length. The solid red trendline depicts the results of a nonlinear one phase-association regression model (dashed lines show 95% confidence intervals). RMSE: root mean squared error, IS: interdaily stability, IV: Intradaily variability, M10: mean activity during most active five hours of the day, L5: mean activity during the least five active hours of the day, DFA: detrended fluctuation analysis.

**Figure 2.**
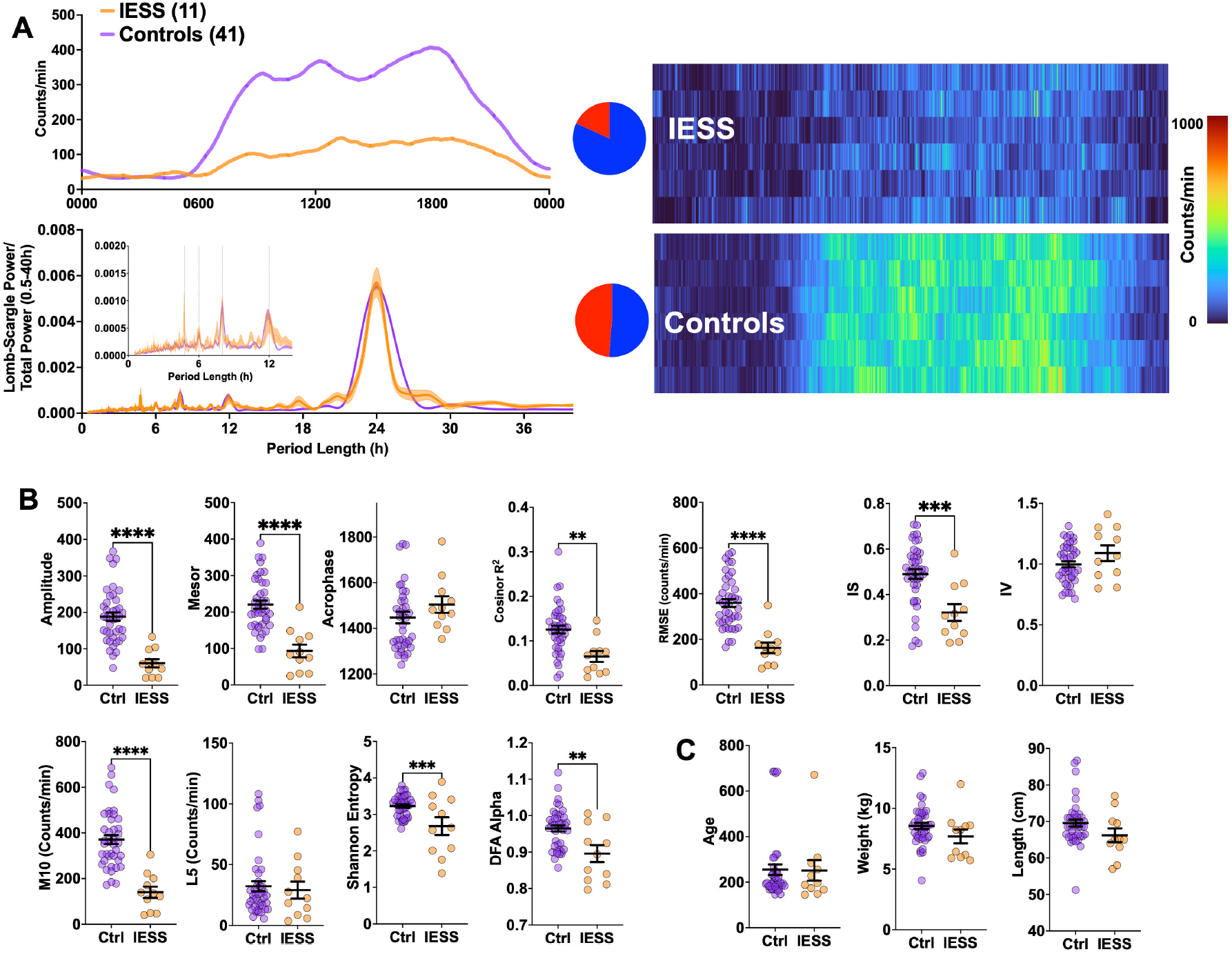
Cross-sectional Comparison of IESS and SHINE Control RARs. A: Smoothed and averaged 24h patterns of rest and activity across groups (top left) with Lomb-Scargle periodograms depicting the relative power of circadian and ultradian peaks across groups (bottom left). RIGHT: Heatmaps depicting averaged (unsmoothed) actigraphy across groups. B: Differences between IESS and control subjects across both parametric and nonparametric RAR measures. C: Age-matched control subjects were not significantly different in average age, weight or length. Mean + standard error of the mean shown. **, *** and **** depict p<0.01, 0.001 and 0.0001 respectively on Student’s t test. RMSE: root mean squared error, IS: interdaily stability, IV: Intradaily variability, M10: mean activity during most active five hours of the day, L5: mean activity during the least five active hours of the day, DFA: detrended fluctuation analysis.

**Figure 3.**
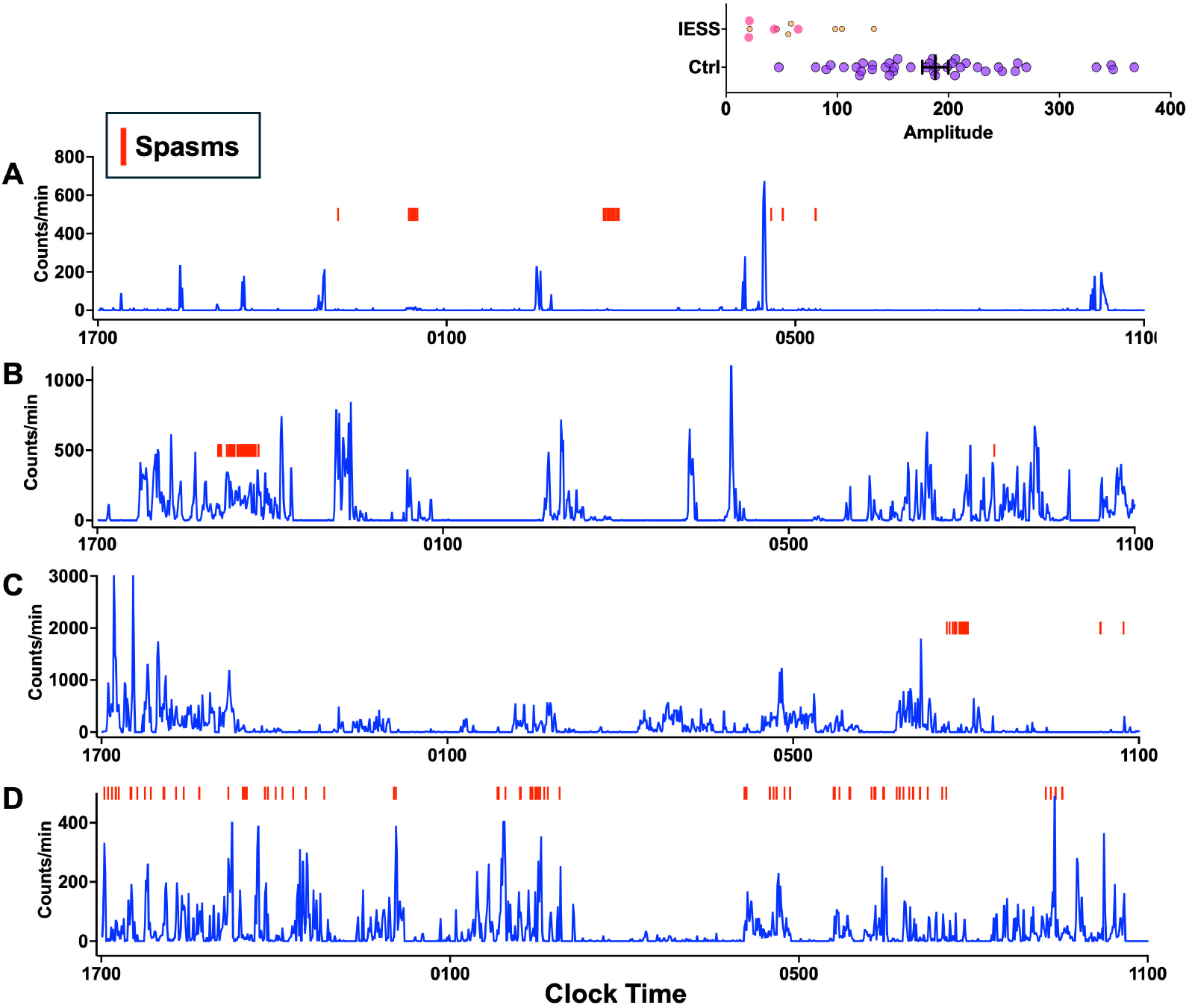
Spasm Occurrence in Relation to Continuous Actigraphy Recordings. A-D: Raw actigraphy recordings (1700 to 1100) for four subjects are shown in relation to the timing of epileptic spasms (red bars), as determined by close review of the video EEG recording. Inset depicts RAR amplitudes for these four patients (pink), in relation to amplitudes for remaining 7 IESS subjects (orange) and age-matched controls (purple)

### RAR Metrics

We devised a single MATLAB function to calculate both parametric and nonparametric RAR measures. Traditional cosinor analyses using least-squares regression was applied to estimate RAR amplitude, mesor (rhythm-adjusted mean) and acrophase (timing of peak activity)^14^. Model R^2^ and root mean square error was also measured to estimate goodness of fit. IV (intradaily variability), IS (interdaily stability), M10 (mean activity during the 10 most active hours of the day) and L5 (mean activity during the least active 5 hours of the day) were calculated as described previously^14,16,25,27,28^. Shannon entropy was calculated by binning activity counts into 100 patient-specific equally spaced bins spanning the observed range of values. The probability distribution of bin occupancy was computed, and entropy as *H* = −∑*p*_*i*_log_2_(*p*_*i*_). Higher values indicating greater behavioral complexity and reduced predictability^45^. Fractal complexity was quantified using detrended fluctuation analysis (DFA)^15^, in which the integrated, mean-centered activity time series was linearly detrended across non-overlapping windows of 60, 360, 7200, 1080 and 1440 minutes. The DFA scaling exponent (alpha) was estimated as the slope of the log-log relationship between fluctuation magnitude and window size. To visualize age-related trends (Fig. 2B), data were fit using a nonlinear one phase-association (rise to plateau) regression model (Prism GraphPad 11). Fitted curves are presented for descriptive purposes only and were not utilized to infer the true functional form of the age-related relationship or specifically test for asymptomatic behavior. 95% confidence intervals are shown to estimate uncertainty in the modeled trajectory, not interindividual variability. Student’s t tests were employed to compare IESS and SHINE controls (Fig. 2). Multiple linear regression was employed to assess associations of age, sex and race/ethnicity with RAR metrics, body weight and length. For each outcome measure, standardized regression coefficients (β) and 95% confidence intervals were visualized using forest plots (Prism GraphPad 11, Fig. S1).

## Results

From a total of 1257 SHINE actigraphy recordings, we excluded those that contained >10% missing values (315) and < 4 full days captured (28). Among 891 included recordings from 333 unique subjects, the median recording duration was 6 days (range 5-13) with a median missingness of 1.43% (0-10%). In Fig. 1A, we illustrate individual actograms from representative SHINE subjects captured during visits 1-4. To visualize overall trends, we provide smoothed and averaged actograms, as well as unsmoothed averaged heatmaps (Fig. 1B). Relatively blunted diurnal fluctuations in activity were present at visit 1. By visit 4, RARs featured a robustly increased height/amplitude together with active periods that exhibited a distinct bimodal profile. We then applied Lomb-Scargle periodograms to estimate the relative spectral power of rhythmic components with periods ranging from 0.5-40h. Circadian rhythmicity (∼24h) was the dominant spectral feature at all four visits, although its relative power was attenuated at visit 1.

Next, we generated X-Y plots to visualize how each RAR metric individually varied with age. To illustrate any potential developmental trends, data were fit to a nonlinear one-phase model. Age-related trajectories differed across metrics, with some exhibiting asymptotic behavior, while others showed no evidence of reaching a plateau. Among parametric/cosinor endpoints, we observed robust age-dependent increases in RAR amplitude, mesor and root mean square error (RMSE), pointing to increases in total activity together with a simultaneous increase in the magnitude of residual variation from the modeled cosine curve. RAR phase (acrophase) and model goodness of fit (R^2^) values plateaued following visit 2 (∼6 months of age), reflecting stabilization of circadian organization and rhythm timing (Fig. 1C). Nonparametric analyses revealed progressive reductions in IV and increases in IS, capturing improvements in day-to-day RAR consistency and within-day RAR activity fragmentation. Changes to M10 mirrored amplitude/mesor changes, while L5 measures did not evolve. RAR entropy measures appeared to stabilize by visit 2, while age-dependent increases in fractal complexity appeared to stabilize by visit 3 (∼1-year olds, Fig. 1D). Body weight and length measurements followed expected developmental trajectories (Fig. 1D). Together, these findings capture a neurodevelopmental trajectory of RAR maturation that occurs in parallel with physical growth.

To examine how age-related changes in RAR metrics, weight and length, were influenced by demographic factors, we modeled each outcome as a function of age, sex and race/ethnicity (comparing Black/African American, Asian or Hispanic subjects to White, Fig. S1). Increasing age was significantly associated with increases in nearly every RAR metric (amplitude, mesor, IS, RMSE/R^2^, M10, α (DFA scaling exponent), Shannon entropy and weight/length. Age displayed a weak negative association with acrophase and strong inverse association with IV. Female sex was associated with more subtle reductions in measures of RAR height (amplitude, mesor, M10) as well as weight/length. Smaller race/ethnicity related differences were seen except in measurements of RAR acrophase, which was significantly delayed in all three classes of non-white subjects. This aligns with previous SHINE results demonstrating the emergence of racial and ethnic differences in objectively measured sleep timing in subjects from visits 1 and 2^38^.

We approached 27 families of patients with IESS. 7 declined to participate and 7 additional participants did not return the actigraphy device. Technical errors limited data collection to a single day in two subjects. We included a total of 11 IESS subjects with multiday actigraphy recordings for RAR analysis. Demographic and treatment data are shown in Table 1 as aggregate measures to preserve participant anonymity. The median recording duration was 10 days (range 4–16), and all included recordings contained no missing data.

**Table 1.**

| **Table 1** |  |
| --- | --- |
| **Median Age [Range]** | 196 [145-671] |
| **Sex** | 9 male, 2 female |
| **Gestational Age at Birth (weeks)** |  |
| 28-31.9 | 2 (18%) |
| 32-36.9 | 2 (18%) |
| ≥37 | 7 (64%) |
| **Treatment** |  |
| Prednisolone | 11 (100%) |
| Vigabatrin | 2 (18%) |
| Levetiracetam | 4 (36%) |
| Valproic acid | 1 (9%) |
| **Persistent Spasms** | 4 (36%) |

In Fig. 2, we provide a qualitative and quantitative comparison of RARs from these IESS subjects and a total of 41 age-matched SHINE controls. Overall, IESS subjects displayed RARs that were severely dampened in height, without differences in the relative power of circadian and ultradian oscillations in activity (Fig. 2A). As shown in Fig. 2B, IESS subjects displayed significant deficits in RAR height (amplitude, mesor, M10), interdaily stability and fractal complexity/entropy, with poorer goodness of fit. We observed a trend for IESS subjects to be underweight (p= 0.13) and shorter in length (p=0.11). Smoothed actograms for each individual case-control comparison are shown in Fig. S2 and revealed a spectrum of RAR disruption. While several subjects displayed relatively preserved diurnal patterning, others displayed a globally flattened profile with minimal day-night differentiation. Together, these results qualitatively and quantitatively clarify the extent of RAR disruptions in a cohort of infants with IESS.

In line with previous estimates from the same institution^46^, 4/11 (36.3%) IESS patients experienced persistent infantile spasms despite treatment. In Fig. 3A-D, we depict the timing of spasms or spasm clusters aligned with actigraphy data. Spasms occurred across a range of activity states, either clustered within discrete portions of the day (A-C) or more broadly distributed across the day-night cycle (D). Visual inspection did not reveal a consistent relationship between seizure occurrence and momentary activity levels, highlighting inter-individual variability in the temporal association between spasms and rest–activity patterns.

## Discussion

By providing an index of emerging circadian behavior, measurements of rest and activity rhythms during early developmental windows provide objective assessments into the maturation of diurnal behavior. In this study, we leveraged a publicly accessible collection of annotated actigraphy recordings from healthy infants to visualize how RARs mature between 30 days to 2 years of age, extending on a previously published report of RAR maturation in 3–12-month-olds^24^. Our results identify robust developmental trajectories during this window of development, demonstrating age-dependent RAR strengthening through increases in amplitude, consistency and complexity. Using the same activity monitor, we identified severely disrupted RARs in a cohort of infants with an EEG-confirmed diagnosis of epileptic spasms. These included deficits in RAR height (amplitude, mesor), robustness/consistency (R^2^, IS) together with weakened temporal complexity and multiscale structure (DFA alpha, Shannon entropy). As with any cross-sectional case-control actigraphic comparison, the etiology and/or stability of these RAR changes is unknown. All 11 patients in our IESS cohort were actively receiving treatment with prednisolone, which can cause irritability and weight gain^47^ as adverse effects, rather than sedation or psychomotor slowing. Low amplitude and variable/inconsistent RARs may reflect the extent of a developmental encephalopathy/lethargy driven by the underlying genetic or structural etiology. Longitudinal prospective actigraphic assessments are necessary to determine whether improvements in RAR structure correlate with measurable gains in cognitive/motor/emotional milestones and both subjective and objective aspects of sleep impairment^34,48^.

Our study has several limitations. First, our cohort size was small, limiting any meaningful subgroup analysis by etiology, treatment or treatment-resistance. Our sample was recruited from a tertial epilepsy center, which may over-represent severe or treatment-resistant cases. In comparison to our previous RAR characterizations in adults with epilepsy^27,28^, we observed a high device non-return rate (∼26%). This may in part be related to the emotional tolls imposed by caring for an infant with epileptic encephalopathy^49^. Second, our IESS cohort primarily sampled male patients (9/11), while our selected controls had a more even sex distribution (21/41 males). In SHINE subjects (Fig. S1), male sex was associated with a higher amplitude, mesor and M10. RARs in IESS subjects displayed significantly depressed amplitudes despite their relative male predominance. Observed differences between IESS and SHINE control subjects may also be linked to differences in climate^50^ (Houston vs Boston). We also did not account for the effects of prematurity, which frequently imposes brain injuries that are linked to IESS^30^. 4/11 IESS subjects were born pre-term, while SHINE only included infants born at term^39,40^. Third, we did not directly measure or compare sleep amount or timing. Algorithms to estimate sleep using actigraphy data alone tend to suffer from poor wake specificity (incorrectly labeling quiet periods of wakefulness as sleep) and are therefore ideally performed in conjunction with sleep diaries^38,39^. To encourage participation and adherence, we deferred the need for sleep diaries. IESS subjects were also not provided a financial reimbursement for their participation. Fourth, actigraphy alone was insufficient to identify the presence or timing of spasms themselves. Future studies designed to employ limb accelerometry as a spasm detector may benefit from more granular measurements made possible through devices that measure high resolution (∼32Hz) triaxial accelerometry.

In conclusion, we show that serially measured rest-activity rhythms derived from ankle actigraphy reveal a infant RAR growth curve, featuring improvements in rhythm height, consistency, complexity and fragmentation. In an etiologically diverse cohort of infants with infantile spasms, we identify marked deficits (or delays) in the same objectively measured parameters. These results lend to future studies that longitudinally appraise the natural history of RARs in IESS and other infantile onset epilepsies, providing guideposts for disease-modifying interventions designed to improve both ictal and interictal aspects of disability.

## Acknowledgements and Conflicts of Interest

We acknowledge Ms. Norma Hernandez, RN (Texas Children’s Hospital) for her role in identifying study subjects. This study did not receive any targeted funding. VK’s laboratory receives funding from the NINDS (R01NS13199). VK is a scientific advisor for Enliten AI and a member of the Digital In Vivo Alliance (DIVA).

## Data Availability statement

SHINE data are publicly available through the National Sleep Research Resource (www.sleepdata.org). De-identified IESS data may be shared with qualified investigators upon reasonable request.

## Authors contributions

VK conceptualized the study. REA, SK, and VK drafted the manuscript. REA, SK, NA, DST acquired clinical data. REA, DST and VK interpreted clinical data. NA, CAN, AGM, and VK performed the analysis and interpretation of data. All authors revised the manuscript and approved content.

---

**Supplemental Figure 1.**
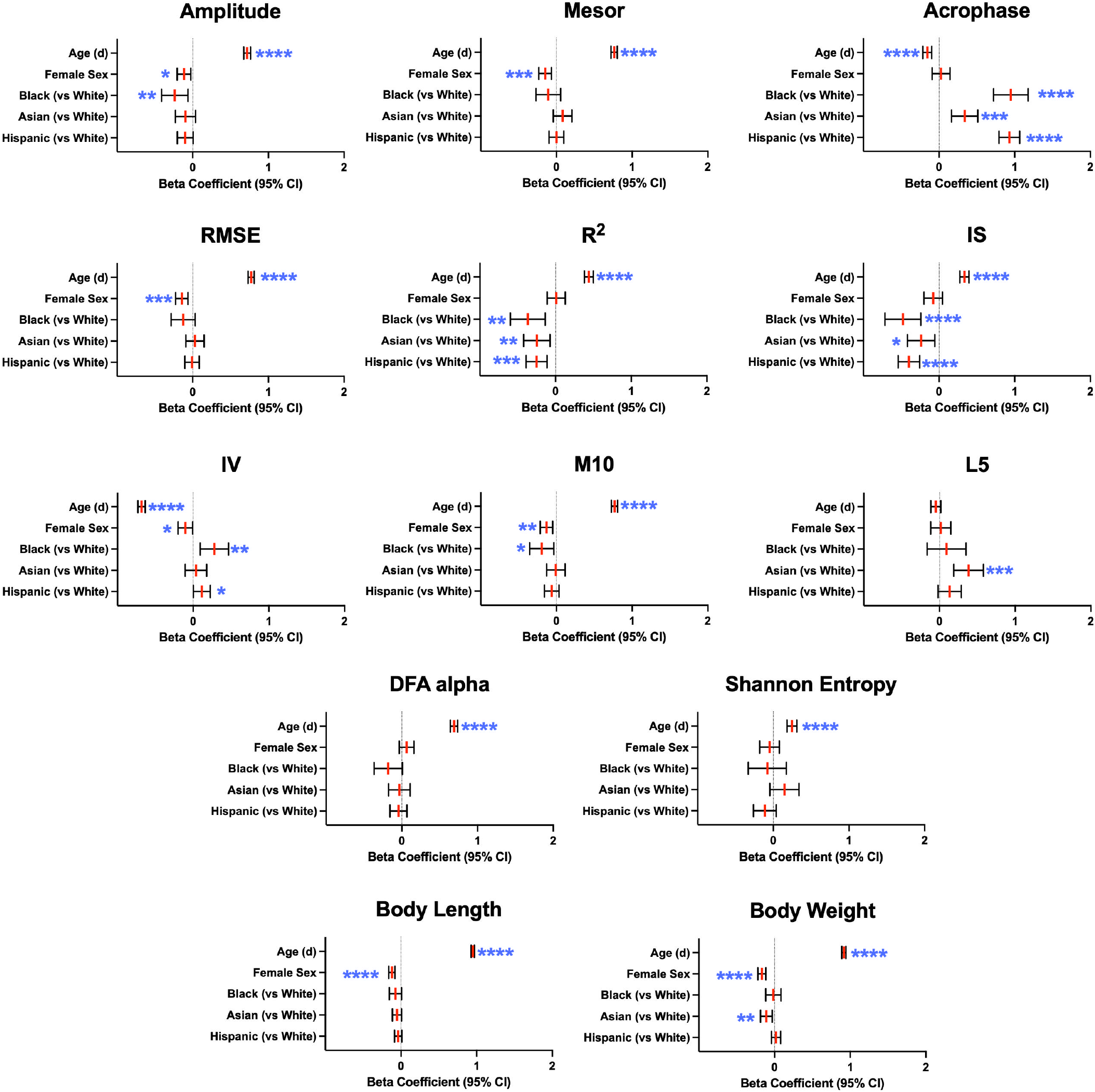
Height, Weight and RAR Metrics Vary by Age, Sex and Racial/Ethnic Differences. Using the SHINE dataset, multiple linear regression models were employed to assess associations of age, sex and race/ethnicity with RAR metrics, body weight and length. Standardized regression coefficients (β) and 95% confidence intervals are shown. *, **, *** and **** depict p<0.05, 0.01, 0.001 and 0.0001 respectively. RMSE: root mean squared error, IS: interdaily stability, IV: intradaily variability, M10: mean activity during most active five hours of the day, L5: mean activity during the least five active hours of the day.

**Supplemental Figure 2.**
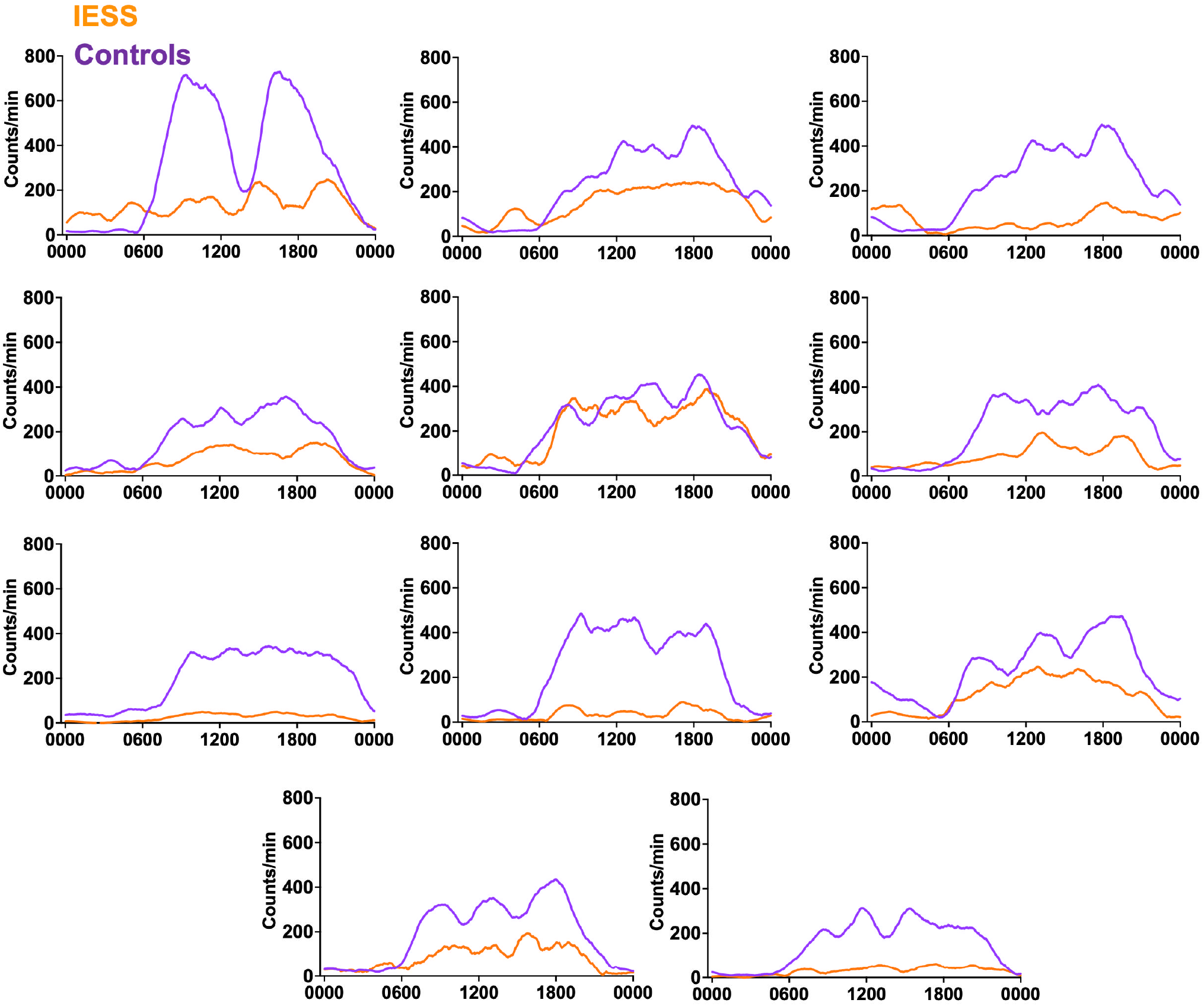
Case Control Actigraphy Comparisons. Smoothed and averaged multiday actograms are shown for each of 11 IESS subjects (orange) and their respective age-matched controls (purple). Curves are presented in a random order.

## References

1. Zubler JM, Wiggins LD, Macias MM, et al. Evidence-Informed Milestones for Developmental Surveillance Tools. Pediatrics. Mar 1 2022;149(3) doi:10.1542/peds.2021-052138

2. Gilmore JH, Knickmeyer RC, Gao W. Imaging structural and functional brain development in early childhood. Nat Rev Neurosci. Feb 16 2018;19(3):123–137. doi:10.1038/nrn.2018.1

3. Miike T. Appropriate Lifelong Circadian Rhythms Are Established During Infancy: A Narrative Review. Clocks Sleep. Aug 7 2025;7(3) doi:10.3390/clockssleep7030041

4. Mirmiran M, Maas YG, Ariagno RL. evelopment of fetal and neonatal sleep and circadian rhythms. Sleep Med Rev. Aug 2003;7(4):321–34. doi:10.1053/smrv.2002.0243

5. Pillai M, James DK, Parker M. The development of ultradian rhythms in the human fetus. Am J Obstet Gynecol. Jul 1992;167(1):172–7. doi:10.1016/s0002-9378(11)91654-8

6. Mendez N, Corvalan F, Halabi D, et al. From gestational chronodisruption to noncommunicable diseases: Pathophysiological mechanisms of programming of adult diseases, and the potential therapeutic role of melatonin. J Pineal Res. Dec 2023;75(4):e12908. doi:10.1111/jpi.12908

7. Rivkees SA. Developing circadian rhythmicity in infants. Pediatr Endocrinol Rev. Sep 2003;1(1):38–45.

8. Blumberg MS, Gall AJ, Todd WD. The development of sleep-wake rhythms and the search for elemental circuits in the infant brain. Behav Neurosci. Jun 2014;128(3):250–63. doi:10.1037/a0035891

9. Zornoza-Moreno M, Fuentes-Hernandez S, Sanchez-Solis M, Rol MA, Larque E, Madrid JA. Assessment of circadian rhythms of both skin temperature and motor activity in infants during the first 6 months of life. Chronobiol Int. May 2011;28(4):330–7. doi:10.3109/07420528.2011.565895

10. O’Connor C, Ventura S, Proietti J, O’Sullivan MP, Boylan GB, Neonatal Sleep Talks g. Sleep and infant development in the first year. Pediatr Res. Feb 2 2026; doi:10.1038/s41390-026-04780-4

11. Halal CSE, Matijasevich A, Howe LD, Santos IS, Barros FC, Nunes ML. Short Sleep Duration in the First Years of Life and Obesity/Overweight at Age 4 Years: A Birth Cohort Study. J Pediatr. Jan 2016;168:99–103 e3. doi:10.1016/j.jpeds.2015.09.074

12. Taveras EM, Rifas-Shiman SL, Oken E, Gunderson EP, Gillman MW. Short sleep duration in infancy and risk of childhood overweight. Arch Pediatr Adolesc Med. Apr 2008;162(4):305–11. doi:10.1001/archpedi.162.4.305

13. Neishabouri A, Nguyen J, Samuelsson J, et al. Quantification of acceleration as activity counts in ActiGraph wearable. Sci Rep. Jul 13 2022;12(1):11958. doi:10.1038/s41598-022-16003-x

14. Krafty RT, Fu H, Graves JL, Bruce SA, Hall MH, Smagula SF. Measuring Variability in Rest-Activity Rhythms from Actigraphy with Application to Characterizing Symptoms of Depression. Stat Biosci. Jul 2019;11(2):314–333.

15. Raichlen DA, Klimentidis YC, Hsu CH, Alexander GE. Fractal Complexity of Daily Physical Activity Patterns Differs With Age Over the Life Span and Is Associated With Mortality in Older Adults. J Gerontol A Biol Sci Med Sci. Aug 16 2019;74(9):1461–1467. doi:10.1093/gerona/gly247

16. Wallace DA, Johnson DA, Redline S, Sofer T, Kossowsky J. Rest-activity rhythms across the lifespan: cross-sectional findings from the US representative National Health and Nutrition Examination Survey. Sleep. Nov 8 2023;46(11) doi:10.1093/sleep/zsad220

17. Li J, Somers VK, Lopez-Jimenez F, Di J, Covassin N. Demographic characteristics associated with circadian rest-activity rhythm patterns: a cross-sectional study. Int J Behav Nutr Phys Act. Aug 18 2021;18(1):107. doi:10.1186/s12966-021-01174-z

18. Li J, Vungarala S, Somers VK D. J, Lopez-Jimenez F, Covassin N. Rest-Activity Rhythm Is Associated With Obesity Phenotypes: A Cross-Sectional Analysis. Front Endocrinol (Lausanne). 2022;13:907360. doi:10.3389/fendo.2022.907360

19. Smagula SF, Zhang G, Gujral S, et al. Association of 24-Hour Activity Pattern Phenotypes With Depression Symptoms and Cognitive Performance in Aging. JAMA Psychiatry. Aug 31 2022; doi:10.1001/jamapsychiatry.2022.2573

20. Kianersi S, Potts KS, Wang H, et al. Chronotype, Life’s Essential 8, and Risk of Cardiovascular Disease: A Prospective Cohort Study in UK Biobank. J Am Heart Assoc. Feb 3 2026;15(3):e044189. doi:10.1161/JAHA.125.044189

21. Musiek ES, Bhimasani M, Zangrilli MA, Morris JC, Holtzman DM, Ju YS. Circadian Rest-Activity Pattern Changes in Aging and Preclinical Alzheimer Disease. JAMA Neurol. May 1 2018;75(5):582–590. doi:10.1001/jamaneurol.2017.4719

22. Nishihara K, Horiuchi S, Eto H, Uchida S. The development of infants’ circadian rest-activity rhythm and mothers’ rhythm. Physiol Behav. Sep 2002;77(1):91–8. doi:10.1016/s0031-9384(02)00846-6

23. Jenni OG, Deboer T, Achermann P. Development of the 24-h rest-activity pattern in human infants. Infant Behav Dev. Apr 2006;29(2):143–52. doi:10.1016/j.infbeh.2005.11.001

24. Rojo-Wissar DM, Bai J, Benjamin-Neelon SE, Wolfson AR, Spira AP. Development of circadian rest-activity rhythms during the first year of life in a racially diverse cohort. Sleep. Jun 13 2022;45(6) doi:10.1093/sleep/zsac078

25. Liguori C, Spanetta M, Fernandes M, Izzi F, Placidi F, Mercuri NB. More than sleep and wake disturbances: An actigraphic study showing the sleep-wake pattern dysregulation in epilepsy. Seizure. Jan 2022;94:95–99. doi:10.1016/j.seizure.2021.11.024

26. Tang T, Zhou Y, Zhai X. Circadian rhythm and epilepsy: a nationally representative cross-sectional study based on actigraphy data. Front Neurol. 2024;15:1496507. doi:10.3389/fneur.2024.1496507

27. Abboud MA, Kamen JL, Bass JS, et al. Actigraphic correlates of neuropsychiatric symptoms in adults with focal epilepsy. Epilepsia. Jun 2023;64(6):1640–1652. doi:10.1111/epi.17611

28. Adhyapak N, Cardenas GE, Abboud MA, Krishnan V. Rest-activity rhythm phenotypes in adults with epilepsy and intellectual disability. Epilepsia Open. Jun 10 2025; doi:10.1002/epi4.70063

29. Meng X, Takacs DS, Kelagere Y, et al. Clinical features of Infantile Epileptic Spasms Syndrome: a systematic review. Orphanet J Rare Dis. Feb 2 2026;21(1) doi:10.1186/s13023-026-04229-1

30. Wheless JW, Gibson PA, Rosbeck KL, et al. Infantile spasms (West syndrome): update and resources for pediatricians and providers to share with parents. BMC Pediatr. Jul 25 2012;12:108. doi:10.1186/1471-2431-12-108

31. Pellock JM, Hrachovy R, Shinnar S, et al. Infantile spasms: a U.S. consensus report. Epilepsia. Oct 2010;51(10):2175–89. doi:10.1111/j.1528-1167.2010.02657.x

32. Bitton JY, Desnous B, Sauerwein HC, et al. Cognitive outcome in children with infantile spasms using a standardized treatment protocol. A five-year longitudinal study. Seizure. Jul 2021;89:73–80. doi:10.1016/j.seizure.2021.04.027

33. Ramgopal S, Shah A, Zarowski M, et al. Diurnal and sleep/wake patterns of epileptic spasms in different age groups. Epilepsia. Jul 2012;53(7):1170–7. doi:10.1111/j.1528-1167.2012.03499.x

34. Hrachovy RA, Frost JD, Jr., Kellaway P. Sleep characteristics in infantile spasms. Neurology. Jun 1981;31(6):688–93. doi:10.1212/wnl.31.6.688

35. Wan L, Yang G, Sun Y, et al. Combined melatonin and adrenocorticotropic hormone treatment attenuates N-methyl-d-aspartate-induced infantile spasms in a rat model by regulating activation of the HPA axis. Neurosci Lett. Mar 23 2021;748:135713. doi:10.1016/j.neulet.2021.135713

36. Wan L, Shi XY, Ge WR, et al. The Instigation of the Associations Between Melatonin, Circadian Genes, and Epileptic Spasms in Infant Rats. Front Neurol. 2020;11:497225. doi:10.3389/fneur.2020.497225

37. Sun Y, Chen J, Shi X, et al. Safety and efficacy of melatonin supplementation as an add-on treatment for infantile epileptic spasms syndrome: A randomized, placebo-controlled, double-blind trial. J Pineal Res. Jan 2024;76(1):e12922. doi:10.1111/jpi.12922

38. Yu X, Quante M, Rueschman M, et al. Emergence of racial/ethnic and socioeconomic differences in objectively measured sleep-wake patterns in early infancy: results of the Rise & SHINE study. Sleep. Mar 12 2021;44(3) doi:10.1093/sleep/zsaa193

39. Quante M, Hong B, von Ash T, et al. Associations between parent-reported and objectively measured sleep duration and timing in infants at age 6 months. Sleep. Apr 9 2021;44(4) doi:10.1093/sleep/zsaa217

40. Quante M, McGee GW, Yu X, et al. Associations of sleep-related behaviors and the sleep environment at infant age one month with sleep patterns in infants five months later. Sleep Med. Jun 2022;94:31–37. doi:10.1016/j.sleep.2022.03.019

41. Li X, Hu H, Quante M, et al. Actigraphy-measured sleep and growth trajectories during the first two years of life: longitudinal evidence from a birth cohort. Sleep. Apr 16 2026;49(4) doi:10.1093/sleep/zsaf386

42. Zhang GQ, Cui L, Mueller R, et al. The National Sleep Research Resource: towards a sleep data commons. J Am Med Inform Assoc. Oct 1 2018;25(10):1351–1358. doi:10.1093/jamia/ocy064

43. Borbely AA, Rusterholz T, Achermann P. Three decades of continuous wrist-activity recording: analysis of sleep duration. J Sleep Res. Apr 2017;26(2):188–194. doi:10.1111/jsr.12492

44. Vieluf S, Cantley S, Krishnan V, Loddenkemper T. Ultradian rhythms in accelerometric and autonomic data vary based on seizure occurrence in paediatric epilepsy patients. Brain Commun. 2024;6(2):fcae034. doi:10.1093/braincomms/fcae034

45. Vagus S, Casey TM, George UZ. Persistent entropy links irregularities in daily and weekly rest and activity cycles during gestation week 22 and 32 to maternal and neonate health outcomes: A prospective cohort study. PLoS One. 2026;21(3):e0342509. doi:10.1371/journal.pone.0342509

46. Katyayan A, Lee ST, Martinez L, Takacs DS. Characteristics of overnight video-EEG monitoring in infantile epileptic spasms syndrome at 2-week follow-up. Epilepsia. Dec 2024;65(12):3583–3594. doi:10.1111/epi.18143

47. Gonzalez-Giraldo E, Stafstrom CE, Stanfield AC, Kossoff EH. Treating Infantile Spasms with High-Dose Oral Corticosteroids: A Retrospective Review of 87 Children. Pediatr Neurol. Oct 2018;87:30–35. doi:10.1016/j.pediatrneurol.2018.06.011

48. Bhanudeep S, Madaan P, Sankhyan N, et al. Long-term epilepsy control, motor function, cognition, sleep and quality of life in children with West syndrome. Epilepsy Res. Jul 2021;173:106629. doi:10.1016/j.eplepsyres.2021.106629

49. Gallop K, Lloyd AJ, Olt J, Marshall J. Impact of developmental and epileptic encephalopathies on caregivers: A literature review. Epilepsy Behav. Oct 1 2021;124:108324. doi:10.1016/j.yebeh.2021.108324

50. Mattingly SM, Grover T, Martinez GJ, et al. The effects of seasons and weather on sleep patterns measured through longitudinal multimodal sensing. NPJ Digit Med. Apr 28 2021;4(1):76. doi:10.1038/s41746-021-00435-2

